# Safety of Breastfeeding in Mothers with SARS-CoV-2 Infection

**DOI:** 10.1101/2020.05.30.20033407

**Authors:** Qingqing Luo, Lan Chen, Dujuan Yao, Jianwen Zhu, Xiangzhi Zeng, Lin Xia, Min Wu, Lin Lin, Zhishan Jin, Qingmiao Zhang, Dilu Feng, Shihuan Yu, Bo Song, Wanlin Zhang, Hongbo Wang, Yanfang Zhang, Zhengyuan Su, Minhua Luo, Xuan Jiang, Hui Chen

## Abstract

**Background:** The outbreak of Coronavirus Disease 2019 (COVID-19) is threatening a surging number of populations worldwide, including women in breastfeeding period. Limited evidence is available concerning breastfeeding in women with COVID-19.

**Methods:** Twenty-three pregnant women and puerperae were enrolled in the study. To evaluate the effect of breastfeeding on SARS-CoV-2 transmission, the presence of SARS-CoV-2, IgG and IgM in breast milk, maternal blood and infant blood were assessed. Feeding patterns were also recorded in follow-up.

**Results:** No positive detection for SARS-CoV-2 of neonates was found. All breast milk samples were negative for the detection of SARS-CoV-2. The presence of IgM of SARS-CoV-2 in breast milk was correlated with maternal blood. The results of IgG detection for SARS-CoV-2 were negative in all breast milk samples. All the infants were in healthy condition while six of them were fed with whole or partial breast milk. Eight infants received antibody test for SARS-CoV-2 in one month after birth and the results were all negative.

**Conclusion:** Findings from this small number of cases suggest that there is currently no evidence for mother-to-child transmission via breast feeding in women with COVID-19 in the third trimester and puerperium.

## Introduction

The outbreak of the 2019 novel coronavirus disease (COVID-19) caused by severe acute respiratory syndrome coronavirus 2 (SARS-CoV-2) was started in Wuhan, China, since December 2019, and rapidly spread to the rest of the country. COVID-19 also occurred in other countries and become a global threat. Confirmed cases of COVID-19 all over the world have surpassed 690 000 with over 33 000 deaths by March 31st, 2020.

The genome of SARS-CoV-2 shares about 80% sequence identity with other two coronaviruses causing emergent public health events in recent decades, severe acute respiratory syndrome coronavirus (SARS-CoV) and Middle East respiratory syndrome virus (MERS-CoV) [1]. The epidemiology of COVID-19 is similar to that of SARS and MERS with droplets and close contact as the main transmission route [2]. Based on the understanding of SARS and MERS, control measures provided by World Health Organization include maintaining distance to avoid close contact. Although fatality rate of SARS-CoV-2 seems to be lower than that of SARS and MERS, more people are threatened by COVID-19 because of much higher infectious capability [3]. Large number of specific populations such as women in the breastfeeding period suffer from the threat of the disease. Previously, a study involved nine pregnant women confirmed with COVID-19 and their infants showed no direct evidence of maternal-infant transmission, but little was reported on how breastfeeding would affect SARS-CoV-2 transmission [4]. Breastfeeding involves a serious of intimate behaviors including skin-to-skin touch and inadvertent cough or sneeze between mothers and infants. Guidelines on breastfeeding in women with COVID-19 are controversial. The guideline for pregnant women suspected with SARS-CoV-2 suggested isolation of mothers from infants without breastfeeding until viral shedding is clear [5] in consideration of possible transmission caused by the intimate behaviors. Similarly, expert consensus from Chinese Obstetricians and Gynecologists Association (COGA) does not recommend breastfeeding [6]. However, there is no enough evidence for mother-to-child transmission (MTCT) up to now. Royal College of Obstetricians & Gynaecologists (RCOG) suggests breastfeeding since they believed the benefits of breastfeeding outweigh the potential risks of MTCT [7]. Therefore, the safety of breastfeeding in women with COVID-19 is worth exploring. In this study, we performed a preliminary study to evaluate the safety of breastfeeding in women infected with SARS-CoV-2.

## Methods

### Study design and patients

Total twenty-three pregnant women with fourteen confirmed with SARS-CoV-2 infection and nine suspected with COVID-19 were enrolled in the study. The confirmation of SARS-CoV-2 infection and the onset of illness took place all in the third trimester and puerperium. All the patients were admitted in Western Branch of Union Hospital, Tongji Medical College, Huazhong University of Science and Technology (HUST) from February 1st to March 15th, 2020. Diagnosis of COVID-19 was based on the New Coronavirus Pneumonia Prevention and Control Program (7th edition) published by the National Health Commission of China [8]. The presence of SARS-CoV-2 nucleic acid in throat swabs or blood was determined by quantitative real-time polymerase chain reaction. The detection of both IgM and IgG for SARS-CoV-2 from break milk or patients’ serum was performed by ELISA. Suspected cases were defined as those with typical changes in chest CT scan of viral pneumonia, but with negative etiologic or serological evidence of SARS-CoV-2 infection. This study was performed according to the Declaration of Helsinki and approved by the Medical Ethical Committee of Union Hospital, Tongji Medical College, HUST (approval number 20200047). Written informed consent forms were obtained from all enrolled patients.

### Data and sample collection

Medical record data of all patients were reviewed. Respiratory tract samples from the patients and throat swabs of their neonates were collected after delivery. Sample collection was done by following the guidelines for COVID-19 issued by Chinese Center for Disease Control and Prevention (CDC). Briefly, clinical specimens from tonsils and posterior pharyngeal wall were collected by synthetic fiber swabs and conserved in sterile tubes containing 3 ml of viral transport medium. Breast milk samples were collected from puerperae and stored in −80. Sample testing was performed in Department of Clinical Laboratory, Union Hospital Tongji Medical College, HUST and Wuhan Institute of Virology, Chinese Academy of Sciences by detection of ORF1ab gene and N gene of SARS-CoV-2 (BioGerm, Shanghai, China) as described previously [4]. The detection of IgG and IgM for SARS-CoV-2 in maternal blood and breast milk was performed using IgG/IgM ELISA kit (Livzon, China) according to the manufacturer’s instruction. The detection of IgG and IgM of SARS-CoV-2 in infants was performed one month after birth by colloidal gold stripes (Livzon, China).

### Statistical analysis

Continuous variables were presented as mean ± standard deviation and compared using the Student’s t-test. Categorical variables were expressed as number (%) and compared using chi-square or Fisher’s exact test as appropriate. *P* value less than 0.05 was considered statistically significant. Statistical analysis was done in SPSS version 21.0.

## Results

Diagnosis for COVID-19 of patients were confirmed in the third trimester (20 in 23, 87%) and puerperal period (3 in 23, 13%). Age range of the patients was 21-40 years with an average of 29.2 ± 4.9 years. The gestational week on delivery was ranged from 34 weeks plus 2 days to 41 weeks plus 3 days. Clinical features of these patients on admission were listed in Table 1.

**Table 1.** Maternal clinical characteristics.

|  | Confirmed group | Suspected group |  |
| --- | --- | --- | --- |
| Characteristic | (N=14) | (N=9) | *P* value |
| **Maternal average age — yr** | 30.5±5.2 | 27.2±3.8 | 0.098 |
| **Contact history — no. (%)** | 6 (42.9) | 0 (0) | 0.048 |
| Environment | 0 | 0 |  |
| infected person | 6 (42.9) | 0 (0) | 0.048 |
| **Clinical symptom — no. (%)** |  |  |  |
| Fever | 10 (71.4) | 1 (11.1) | 0.009 |
| Cough | 9 (64.3) | 2 (22.2) | 0.089 |
| Myalgia | 3 (21.4) | 0 | 0.253 |
| Dyspnea | 4 (28.6) | 0 | 0.127 |
| Diarrhea | 1 (7.1) | 0 | 0.609 |
| Asymptomatic | 2 (14.3) | 6 (66.7) | 0.023 |
| **Delivery mode — no. (%)** |  |  |  |
| Cesarean section | 12 (85.7) | 5 (55.6) | 0.162 |
| Vaginal delivery | 2 (14.3) | 4 (44.4) | 0.162 |
| **Maternal death** — no. (%) | 0 | 0 |  |
| **Antiviral therapy** — no. (%) |  |  |  |
| Antepartum | 6 (42.9 ) | 2 (22.2) | 0.4 |
| Postpartum | 10 (71.4) | 4 (44.4) | 0.196 |
| **Glucocorticoid therapy** — no. (%) |  |  |  |
| Antepartum | 6 (42.9) | 1 (11.1) | 0.106 |
| Postpartum | 10 (71.4) | 3 (33.3) | 0.72 |

None of the patients reported to have direct contact with Huanan Wholesale Seafood Market. In total, six patients who were all from the confirmed group reported that they had a clear contact history with confirmed COVID-19 patients. Fever was the most common complaint when patients were admitted, which had a significantly higher rate in confirmed group than in suspected group (71.4% vs 11.1%, *p*<0.05). Other symptoms of an upper respiratory tract infection were also reported without statistical difference: eleven patients with cough (nine in confirmed group and two in suspected group), three with myalgia (all from confirmed group) and two with dyspnea (all from confirmed group). One patient presented diarrhea from confirmed group, which is an atypical symptom of COVID-19. Notably, eight patients were asymptomatic, and the rate of asymptomatic patients was significantly lower in confirmed group than that in suspected group (14.3% vs 55.6%, *p*<0.05).

Most cases were terminated with cesarean section. None of the patients required mechanical ventilation. Six (42.9%) confirmed patients and 2 (22.2%) suspected patients received antepartum antiviral therapy. Ten (71.4%) confirmed patients and four (44.4%) suspected patients received antiviral treatment after delivery. Six (42.9%) confirmed patients and one (11.1%) suspected patients received antepartum glucocorticoid therapy. Ten (71.4%) confirmed and three (33.3%) suspected patients received hormone after delivery. There were no statistical differences in these treatments between the two groups.

All the pregnancies were singleton. The average birth weight was 3173.9 ± 470.7g with no clinical manifestation of neonatal asphyxia. SARS-CoV-2 detection of throat swab was performed in fifteen neonates at birth and in six neonates in neonatal intensive care unit (NICU) after birth. All the results of SARS-CoV-2 testing in neonates were negative. Clinical features of neonates were displayed in Table 2. Feeding patterns and health conditions of the infants were followed by March 27, 2020 (Table 2). All infants in confirmed group were discharged from NICU, and had no pneumonia-related symptom. Six infants were fed with whole or partial breast milk. Eight infants (five in confirmed group and three in suspected group) received antibody testing one month after birth. The testing was not performed in other fifteen infants because it was rejected by their parents or the infants were under one month. The results of IgM or IgG detection in infants were all negative (Table 2).

**Table 2.**
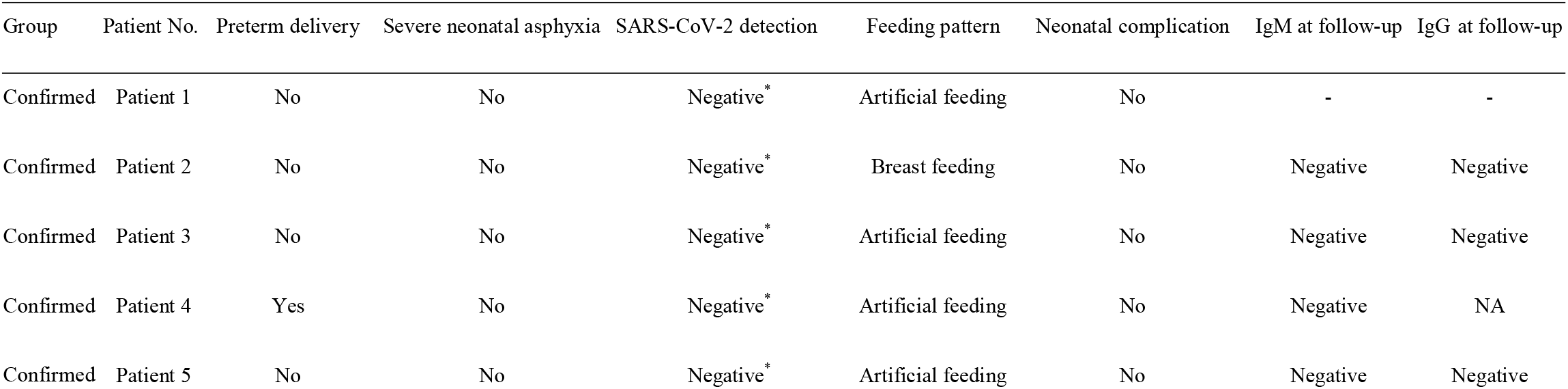
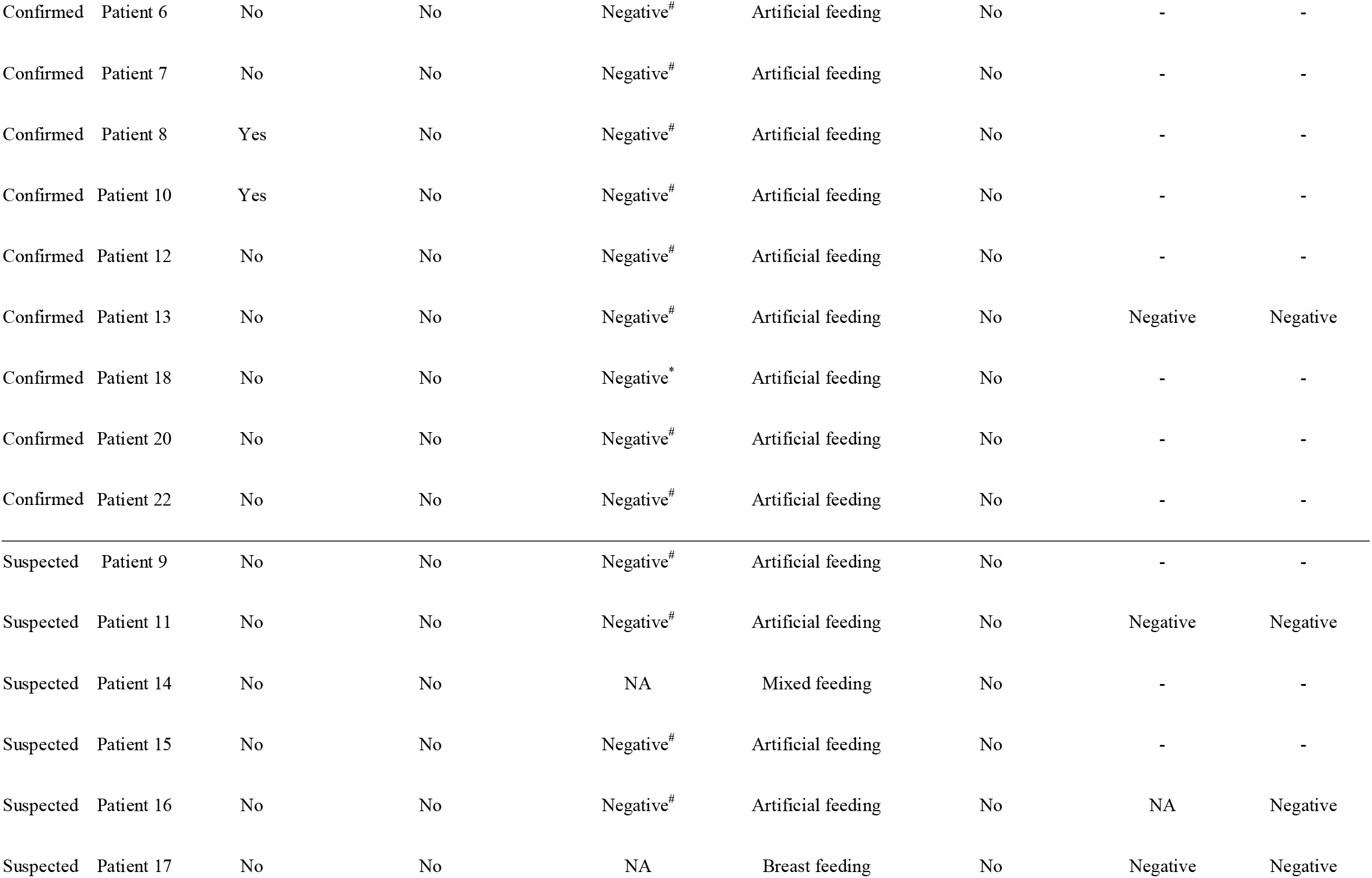
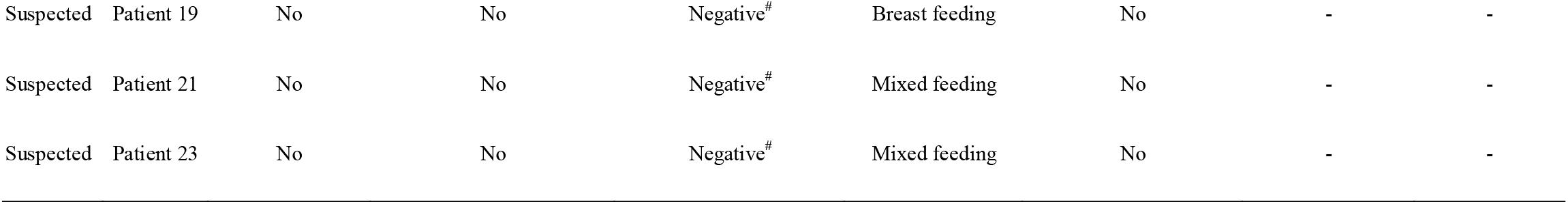
Neonatal outcomes and follow-up. *: throat swab performed in neonatal intensive care unit; #: throat swab performed at birth; NA: not applicable

Most of the breast milk samples were collected within one week postpartum. Samples from two patients were collected on the 15th and 12th day after delivery, respectively, since the puerperae were diagnosed as COVID-19 after delivery. In confirmed group, six samples were collected at the time when SARS-CoV-2 detection in throat swab became negative. The results of SARS-CoV-2 nucleic acid detection in all breast milk samples were negative (Table 3). Testing of IgG and IgM for SARS-CoV-2 in breast milk and maternal blood was performed in seven patients (four confirmed and three suspected) (Table 4). IgM antibody was present in all four confirmed patients (100%) and one suspected patient (33.3%), which was in accordance with IgM detection in the maternal blood. IgG antibody was all negative whatever it was positive or negative in maternal blood (Table 4).

**Table 3.** SARS-CoV-2 detection in breast milk and throat swab.

| Group | Patient No. | Onset of illness to delivery (d) | Sampling day after delivery (d) | SARS-CoV-2 detection in breast milk | Throat swab of SARS-CoV-2 detection when sampling |
| --- | --- | --- | --- | --- | --- |
| Confirmed | Patient 1 | After delivery | 15 | Negative | Negative |
| Confirmed | Patient 2 | Asymptomatic | 7 | Negative | Positive |
| Confirmed | Patient 3 | 13 | 7 | Negative | Positive |
| Confirmed | Patient 4 | 15 | 7 | Negative | Positive |
| Confirmed | Patient 5 | 14 | 5 | Negative | Positive |
| Confirmed | Patient 6 | 16 | 4 | Negative | Positive |
| Confirmed | Patient 7 | 8 | 5 | Negative | Positive |
| Confirmed | Patient 8 | 21 | 3 | Negative | Positive |
| Confirmed | Patient 10 | 9 | 1 | Negative | Positive |
| Confirmed | Patient 12 | 21 | 3 | Negative | Negative |
| Confirmed | Patient 13 | 10 | 3 | Negative | Negative |
| Confirmed | Patient 18 | After delivery | 12 | Negative | Negative |
| Confirmed | Patient 20 | 42 | 3 | Negative | Negative |
| Confirmed | Patient 22 | Asymptomatic | 2 | Negative | Negative |
| Suspected | Patient 9 | 2 | 2 | Negative | Negative |
| Suspected | Patient 11 | 4 | 1 | Negative | Negative |
| Suspected | Patient 14 | After delivery | 5 | Negative | Negative |
| Suspected | Patient 15 | Asymptomatic | 4 | Negative | Negative |
| Suspected | Patient 16 | Asymptomatic | 5 | Negative | Negative |
| Suspected | Patient 17 | Asymptomatic | 6 | Negative | Negative |
| Suspected | Patient 19 | Asymptomatic | 7 | Negative | Negative |
| Suspected | Patient 21 | Asymptomatic | 4 | Negative | Negative |
| Suspected | Patient 23 | Asymptomatic | 4 | Negative | Negative |

**Table 4.** Antibody detection of SARS-CoV-2 IgG and IgM in both breast milk and maternal blood. ±: weakly positive; −: negative; +: positive; NA: not applicable.

|  | Patient 13 | Patient 14 | Patient 15 | Patient 16 | Patient 18 | Patient 20 | Patient 22 |
| --- | --- | --- | --- | --- | --- | --- | --- |
| **Group** | Confirmed | Suspected | Suspected | Suspected | Confirmed | Confirmed | Confirmed |
| **Breast milk** |  |  |  |  |  |  |  |
| IgM | ± | - | ± | - | + | + | ± |
| IgG | - | - | - | - | - | - | - |
| **Blood (before delivery)** |  |  |  |  |  |  |  |
| IgM | + | NA | NA | NA | NA | + | + |
| IgG | + | NA | NA | NA | NA | + | + |
| **Blood (after delivery)** |  |  |  |  |  |  |  |
| IgM | + | NA | ± | - | + | + | + |
| IgG | + | NA | - | - | - | + | + |

## Discussion

In this study, we reported a cohort of twenty-three puerperae confirmed or suspected with COVID-19. The clinical features of these patients showed similar patterns with other patients reported [3]. Fever and cough were the most common symptoms. Gastrointestinal symptom was rare but reported in our cohort. It has been suggested that pregnant women and puerparae within one month after delivery are at greatest risk for respiratory infectious disease like influenza [9]. Cohort studies from SARS and MERS reveal high mortality and increased requirement for mechanical ventilation and ICU admission even when the termination of pregnancy was performed [10, 11]. Severe patients with multiple organ dysfunction syndrome (MODS) and high incidence of severe neonatal asphyxia were reported in pregnancy with COVID-19 [12]. Pregnant outcomes were mild in our study without maternal or fetal/neonatal death, and the medical conditions of the puerperae were stable, which were consistent with the recent reported studies [4, 13].

Human-to-human transmission of SARS-CoV-2 occurred mainly via respiratory droplets [2]. Primary epidemiological feature of COVID-19 is familial cluster [14]. Unlike early reported cases, proportion of patients with contact history of Huanan Wholesale Seafood Market or intimate interactions with confirmed patients was much smaller [4]. In consideration of these information, special attention should be paid to asymptotic case in our cohort. These patient were screened out by chest CT scan before their admissions in other hospitals, and two of them were confirmed as COVID-19 via throat swab specimen detection. Since SARS-CoV-2 is able to be transmitted by asymptomatic carriers [15], routine screening of COVID-19 should be performed before planned admission of pregnant women during epidemic situation for the safety of other inpatients and health care workers. Compared with nucleic acid testing, chest CT scan is more convenient and feasible in most medical institutions with a relative high sensitivity in distinguishing COVID-19 from viral pneumonia [16]. Therefore, chest CT scan should be considered as a primary screening tool for COVID-19 detection before admission of pregnant women. Although the radiation exposure through single examination is much lower than estimated dose for fetus harm, chest CT scan in indicated pregnant patients should following As Low As Reasonably Achievable (ALARA) principle [17].

With extensive progress in the study of SARS-CoV-2, the presence of the virus was verified in patients’ body fluids such as urine and blood besides specimens from respiratory tract [18]. Thus, other possible transmission routes are worth exploring. Presence of viral nucleic acids of SARS-CoV-2 detected in fecal samples points out the probability of fecal-oral transmission in COVID-19 [19]. Negative results of SARS-CoV-2 detection in amniotic fluid and umbilical cord blood indicate a less probability of intrauterine transmission from mother to fetus [4]. However, recent researches reported IgM and IgG antibodies for SARS-CoV-2 were detected in some newborns to mothers with COVID-19 [20], implying the occurrence of *in utero* infection. So far, no literature has described or discussed about the possibility of MTCT via breastfeeding.

Breastfeeding contributes to the health and well-being of both mothers and infants. Early initiation of breastfeeding protects the newborns from acquiring infection and reduces infant mortality [21], especially in premature infants [22]. WHO suggests breastfeeding should be started within one hour after birth and extended until two years old. However, the recommendation becomes complicating when the mothers suffer from infectious diseases. On one hand, immunomodulatory proteins in human milk provide anti-infective benefits for infants, particularly protection against virus causing respiratory and gastrointestinal tract diseases [23]. On the other hand, breast milk may be contaminated with virus such as human cytomegalovirus (HCMV), hepatitis B virus (HBV) and human immunodeficiency virus (HIV), indicating possible risk of MTCT via breastfeeding [24]. Numerous studies have been done in the field of investigating breastfeeding in mothers with viral infection, and the safety of breastfeeding has been reported that no additional risk of MTCT in mothers with HBV or HIV [25, 26]. Continuation of breastfeeding is also recommended in mothers infected with H1N1 influenza [27]. Therefore, breastfeeding should not be simply forbidden in mothers infected with SARS-CoV-2 unless potential risks overweigh advantages of breast feeding.

The absence of SARS-CoV-2 in all breast milk samples in our study was in accordance with the report by Chen et al where breast milk samples were tested from six patients confirmed with COVID-19 [4]. Additionally, our study verified that free of SARS-CoV-2 in breast milk no matter the presence of SARS-CoV-2 was positive or negative in specimens from upper respiratory tract. Even with a small number of cases, this result strongly indicates that breast milk may not be a transmitting vector for SARS-CoV-2, thus breastfeeding is possible for mothers with COVID-19 theoretically. Furthermore, due to the ability of IgG to cross the placenta, the presence of IgG for SARS-CoV-2 in blood samples of mothers before delivery imply the possibility of antepartum protection for fetus. The presence of IgM for SARS-CoV-2 in maternal blood samples was consistent with the presence of IgG. IgG and IgM in breast milk are produced by different mechanisms. Antibody level of IgG and IgM in breast milk were lower than that in maternal blood [28]. IgM antibody of infectious diseases in breast milk is able to provide protection of the same pathogens to infants[29] and inhibit entry and transport of virus such as HIV to infants[30]. We inferred that postpartum protection by antibodies may also exist in SARS-CoV-2 infected mothers to their infants via breastfeeding.

All the infants were in healthy physical conditions by the end of follow-up (March 27, 2020). The rate of breast/mixed feeding was much lower in confirmed group than that in suspected group according to our follow-up. Only one confirmed patient is giving breastfeeding to her infant. The less occasion of breastfeeding in mothers with COVID-19 is due to the cautious suggestion given by COGA under the circumstances that the safety for breastfeeding was uncertain. Besides this case in the study, another patient with confirmed COVID-19 discharged from our hospital has started breastfeeding from middle of February, and no clinical manifestation of pneumonia was developed in her infant (data not shown). The patient was not included in this cohort because we failed to collected her breast milk during her hospitalization. However, due to the lack of evidences from larger sample size in breastfeeding practice, mothers with COVID-19 who have the intention of breastfeeding should be informed of current dilemma of breastfeeding with COVID-19 and possibility of transmission by close contact. Several measures can be performed to limit viral spread as RCOG recommends, such as hand washing before feeding the infants, avoiding coughing or sneezing during feeding and wearing a face mask. Expressed milk for bottle feeding by uninfected relatives may also be an alternative. Furthermore, antiviral remedy should be reconsidered if the mothers are willing to start or continue breastfeeding after confirmed or suspected with COVID-19.

In summary, our study proposed for the first time the feasibility of breastfeeding in women infected with SARS-CoV-2. Taking the potential benefits and risks into account, breastfeeding is encouraged if there is no other medical contradiction. The study was preliminary with a small sample size and short interval for medical observation. The safety of breastfeeding should be proved by further study.

## Data Availability

With the permission of the corresponding authors, clinical data can be provided after publication, without names and identifiers, but not the study protocol, statistical analysis or informed consent form. Research team will provide an email address for communication. The corresponding authors have the right to decide whether to share the data or not.

## Acknowledgements

We thank all the patients involved in the study and their relatives; the midwives, nurses and staffs providing care for the patient; doctors from pediatrics, respiratory medicine, anesthesia and intensive care unit from Union Hospital, HUST and hospitals affiliated of Harbin Medical University.

## Corresponding author contact information

Hui Chen: Department of Obstetrics and Gynecology, Union Hospital, Tongji Medical College, Huazhong University of Science and Technology, 1277 Jiefang Avenue, Wuhan, China 430030.

Xuan Jiang: The Joint Center of Translational Precision Medicine, Guangzhou Institute of Pediatrics, Guangzhou Women and Children Medical Center, Guangzhou, China; The Joint Center of Translational Precision Medicine, Wuhan Institute of Virology, Chinese Academy of Sciences, Wuhan, China 430071.

